# Neural mass modelling reveals that hyperexcitability underpins slow-wave sleep changes in children with epilepsy

**DOI:** 10.1101/2024.07.15.24310128

**Authors:** Dominic M Dunstan, Samantha YS Chan, Marc Goodfellow

## Abstract

**Objective:** The relationship between sleep and epilepsy is important but imperfectly understood. We sought to understand why children with epilepsy have altered sleep homeostasis.

**Methods:** We used neural mass models to replicate sleep EEG recorded from 15 children with focal lesional epilepsies and 16 healthy age-matched controls.

**Results:** The models revealed that sleep EEG differences are driven by enhanced firing rates in the neuronal populations of patients, which arise predominantly due to enhanced excitatory synaptic currents. These differences were more marked in patients who had seizures within 72 hours after the sleep recording. Furthermore, models inferred from patients resided closer in parameter space to models of a typical seizure rhythm.

**Significance:** These results demonstrate that brain mechanisms relating to epilepsy manifest in the interictal EEG in slow-wave sleep, and that EEG recorded from patients can be mapped to synaptic deficits that may explain their predisposition to seizures. Neural mass models inferred from sleep EEG data have the potential to generate new biomarkers to predict seizure occurrence or inform treatment decisions.

**Key Points:**

- The mechanisms that differentiate children with epilepsy from controls during slow-wave sleep can be understood using a mathematical model.
- The observed spectral power shifts in patients are predominately explained by greater excitatory synaptic currents.
- These differences in currents place patients’ models closer to seizure rhythms.
- Ultimately, this framework could help foster the development of biomarkers to guide intervention in epilepsy.

## 3. Introduction

The electroencephalogram (EEG) in deep sleep is dominated by slow waves of high amplitude, generated by widespread neurons alternating in synchrony between depolarised and hyperpolarised states [1]. This slow-wave activity (SWA) correlates closely with sleep need, building up with time spent awake and dissipating with sleep [2]. It has been proposed that the decrease in global SWA across the night reflects the process of synaptic renormalisation [3], a homeostatic mechanism which regulates cortical excitability [4] and facilitates neural plasticity [5]. Further, it is theorised that the disruption of sleep homeostasis may be the basis of sleep-related epilepsies with childhood onset, ranging from self-limited childhood focal epilepsies to mesial temporal lobe epilepsy [6]. In support of this notion, Eriksson et al [7] recently demonstrated that SWA dynamics are altered in children with epilepsy compared to age-matched healthy controls. Moreover, these differences in SWA were found to be exacerbated in patients with a higher propensity to seizures [7].

Understanding the physiological mechanisms that contribute to differences observed in the EEG in disease is a challenging problem [8, 9]. Computational models can help: if we can match the simulations generated by models to data recorded from human subjects, we can then examine which components, parameters or settings of the model were crucial to allow it to generate the data of interest. Several models of EEG have been developed and have been shown to generate dynamics similar to a variety of resting and pathological states [10]. In particular, neural mass models parsimoniously capture synaptic interactions between populations of excitatory and inhibitory neurons [11]. This allows for EEG brain rhythms to be understood in terms of synaptic dynamics, connectivity and firing rates at the level of brain tissue [10, 12, 13, 14].

An important technical challenge is how to match model dynamics to data. Various methods are available for this, including routines within dynamic causal modelling [15]. Such approaches often use linear models [16], combined with prior beliefs on parameter values. The latter are difficult to ascertain for neural mass models, whilst the former does not capture the nonlinear dynamics of the brain and, in particular, the nonlinear mechanisms of epileptiform EEG [17]. Recently, we have demonstrated the promise of multiobjective optimisation as a global nonlinear approach that can be used to model resting and pathological EEG [18]. This method enables the efficient search of parameter space to identify parameter values in the model that, when simulated, produce an output that recapitulates desired features of the data. This process can be applied to data from patients and healthy controls to understand the mechanisms (neural mass model parameters) that are responsible for changes observed in the EEG. Here, we apply this approach to decipher the mechanisms in the sleeping cortex [19] that contribute to the differences in SWA observed in patients with epilepsy. We then explore perturbations to the model that could potentially rectify the different dynamics observed in epilepsy. Finally, having identified mechanisms underpinning differences in resting EEG, we link these mechanisms to the generation of seizures. Here we do this by quantifying the proximity (in terms of similarity of recovered parameter values) of resting dynamics to dynamics of an archetypal seizure rhythm. That is, we answer the questions: “Why would patients have altered SWA?” and “Can this be normalised?”

## 4. Materials and Methods

### 4.1. Participants and study design

Participants consisted of 15 children with drug-resistant focal structural (or presumed structural) aetiology, and 16 age and sex-matched typically developing controls. Patients were recruited prospectively from the EEG video telemetry unit at Great Ormond Street Hospital as described previously [7, 20]. Children were aged between 6 and 16 years, and EEG was recorded continuously during planned four-night hospital admissions. Control participants were recruited by advertisements directed at staff working at the UK charity Young Epilepsy. Controls attended the EEG department of Young Epilepsy to be set up for a single night ambulatory sleep study. Compared to the previous cohort ([7]), 4 patients and 2 controls were excluded due to artefacts in the recordings. Group differences in demographic and clinical data were examined using independent samples t-tests for continuous variables and chi-square tests for categorical variables.

### 4.2. EEG data

EEG polysomnography acquisition, visual sleep scoring and the visual quantification and marking of seizures have been described in previous work [20]. EEG data were recorded with the Xltek Trex system (Natus Medical Incorporated, Pleasanton, CA, USA) at 512Hz using a 10-10 montage (Fz, Cz, Pz, Fp1, Fp2, F3, F4, F7, F8, F9, F10, C3, C4, C5, C6, T5, T6, T7, T8, T9, T10, P3, P4, P9, P10, O1, O2) in patients and at 256Hz using an 8 electrode montage (F3, F4, C3, C4, O1, O2, A1, A2) in controls. Recordings were downsampled to 128Hz in Natus Sleepworks (Natus, USA) before exporting as a .edf file for offline analysis. The full EEG recording (all available channels) was reviewed visually for artefacts in EDFbrowser (version 1.88 https://www.teuniz.net/edfbrowser/), and channels marred by artefacts were excluded. No artefact removal was performed.

To identify early night SWA, the recordings were viewed on a whole night timescale with colour density spectral array to identify segments with high amounts of 1-4Hz with or preceded by high amounts of 10-12 Hz occurring within the first two hours of sleep. The identified segments were reviewed again at a 10-second per-page scale with all channels for visual identification of SWA before cropping. The mean activity recorded from the frontal electrodes (F3, F4) was used for further analysis.

### 4.3. Parent-rated sleep disturbance

Parents were asked to rate the frequency of various sleep behaviours as they would occur in a typical week using the Children’s Sleep Habits Questionnaire [21].

### 4.4. Modelling framework

A neural mass model was used to simulate the temporal dynamics of mean membrane potentials and firing rates in a cortical region [22]. Specifically, we used a conductance-based neural mass model developed to model non-rapid eye movement sleep EEG by Weigenand et al [19]. Here, neurons are grouped into interacting excitatory and inhibitory populations, and excitatory, inhibitory, leak and sodium-dependent potassium (KNa) synaptic currents (at the level of the neural mass) are tracked over time. We note that excitatory synaptic currents exhibit efferent depolarization (labelled as AMPA in [19]) and inhibitory synaptic currents exhibit efferent hyperpolarization (labelled as GABA in [19]). Furthermore, sodium-dependent potassium synaptic currents have been suggested as a mechanism for slow oscillations [19, 23]. We used MATLAB [24] to numerically solve the model. Additional details of the model, including model equations, can be found in Supplemental 1.

The model comprises 32 parameters. These parameters describe the mean synaptic interactions between excitatory and inhibitory neuronal populations. These facilitate a mechanistic interpretation of the neuronal activity analogous to an EEG recording, without explicitly modelling the activity generated by single cells [10, 11]. However, parameters at this scale are difficult to constrain and simulating the model with different parameter values results in different dynamics. Therefore, we implemented a previously developed multiobjective genetic algorithm [18] to search model parameter space for combinations of parameters that yield model simulations with properties similar to those of the EEG. Supplemental Table 1 defines the model parameters, gives a brief description of their interpretation and defines the lower and upper parameter bounds that form the search space.

To quantitatively compare model output with data, objectives that describe the difference between the model and data were defined, and the algorithm was used to recover solutions that minimised these objectives. Following our previous work [18], we defined two objectives: (1) the difference in normalised power; and (2) the difference in node degree of the horizontal visibility graph. The latter objective maps the EEG time series to a network and has been shown to be able to distinguish between stochastic processes and nonlinear dynamics, including epileptiform rhythms [18, 25, 26]. The optimisation was repeated 100 times for each subject to obtain a distribution of parameters that could describe the subject’s EEG data. Figure 1 provides an overview of how parameters were recovered by comparing the model output to the recorded EEG. For further details regarding the definition of objectives, and information on how the model output is aligned to data, see Supplemental 2.

**Figure 1:**
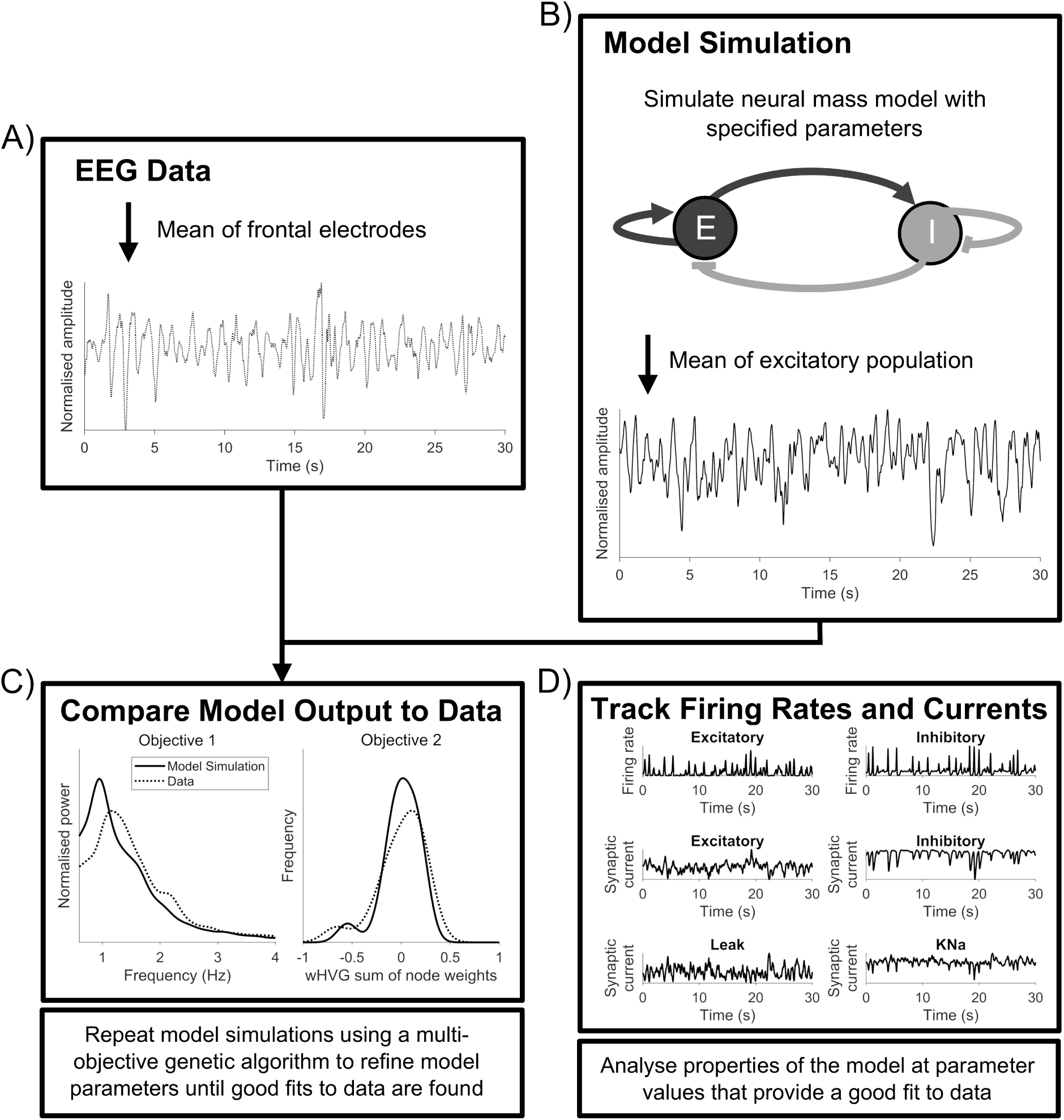
Illustration of the process of comparing model output with data to obtain parameters that explain the EEG. A) The mean signal from the F3 and F4 electrodes of an EEG recording is used in the analysis. Note that a figure of an example patient EEG recording during sleep has been removed here due to potentially identifying information. B) Schematic of the neural mass model used, consisting of interacting excitatory and inhibitory populations of neurons. The output of the model is a 30s time series of simulated EEG. C) Model output is compared to the data by defining objectives to recapitulate in the simulations. Model parameters are adjusted to find simulations that aim to minimise the objectives (see Methods). D) Several properties of the neural mass model are analysed, including firing rates and synaptic currents.

### 4.5. Comparison of resting activity to seizure activity

We additionally optimised the model to generate spike-wave discharges (SWDs). The SWD is the archetypal waveform seen on surface EEG during a generalised seizure [27] and is thought to have a cortical origin [28]. SWDs are activated in sleep across a spectrum of common childhood epilepsies, such as self-limited epilepsy with centrotemporal spikes [29] and various genetic generalised epilepsies [30], as well as in continuous spike-wave during sleep, where SWA is largely replaced by SWD [31]. In animal models, SWDs have been observed to develop from sleep-like slow rhythms [32]. Optimising to this rhythm therefore enabled us to compare (*in silico*) how the mechanisms of a resting state relate to mechanisms of a seizure state.

### 4.6. In silico prediction for intervention

To understand how intervention may alter the dynamics observed on the EEG, we investigated the sensitivity of the power spectrum to changes in model parameters. In particular, we recorded the simulated power spectrum after individually adjusting the parameters governing each synaptic channel conductance (excitatory, inhibitory, leak and KNa). We focused on perturbing the conductance parameters because these parameters represent a key mechanistic target of antiseizure medications (for example, see [33]). This includes benzodiazepines, which have been shown to increase the conductance of GABA_A_ (inhibitory) channels [34] and perampanel, which is known to block AMPA (excitatory) mediated synaptic conductances [35]. Hence, to simulate the effects of treatment, we decreased the excitatory and leak conductances and increased the inhibitory and KNa conductances in models derived from patient data.

## 5. Results

### 5.1. Clinical characteristics of participants

Participant demographics and patient clinical characteristics are summarised in Table 1. In particular, no significant differences in age or sex were recorded between the cohorts. Further information on the epilepsy characteristics of patients is provided in Supplemental Table 2.

**Table 1:**
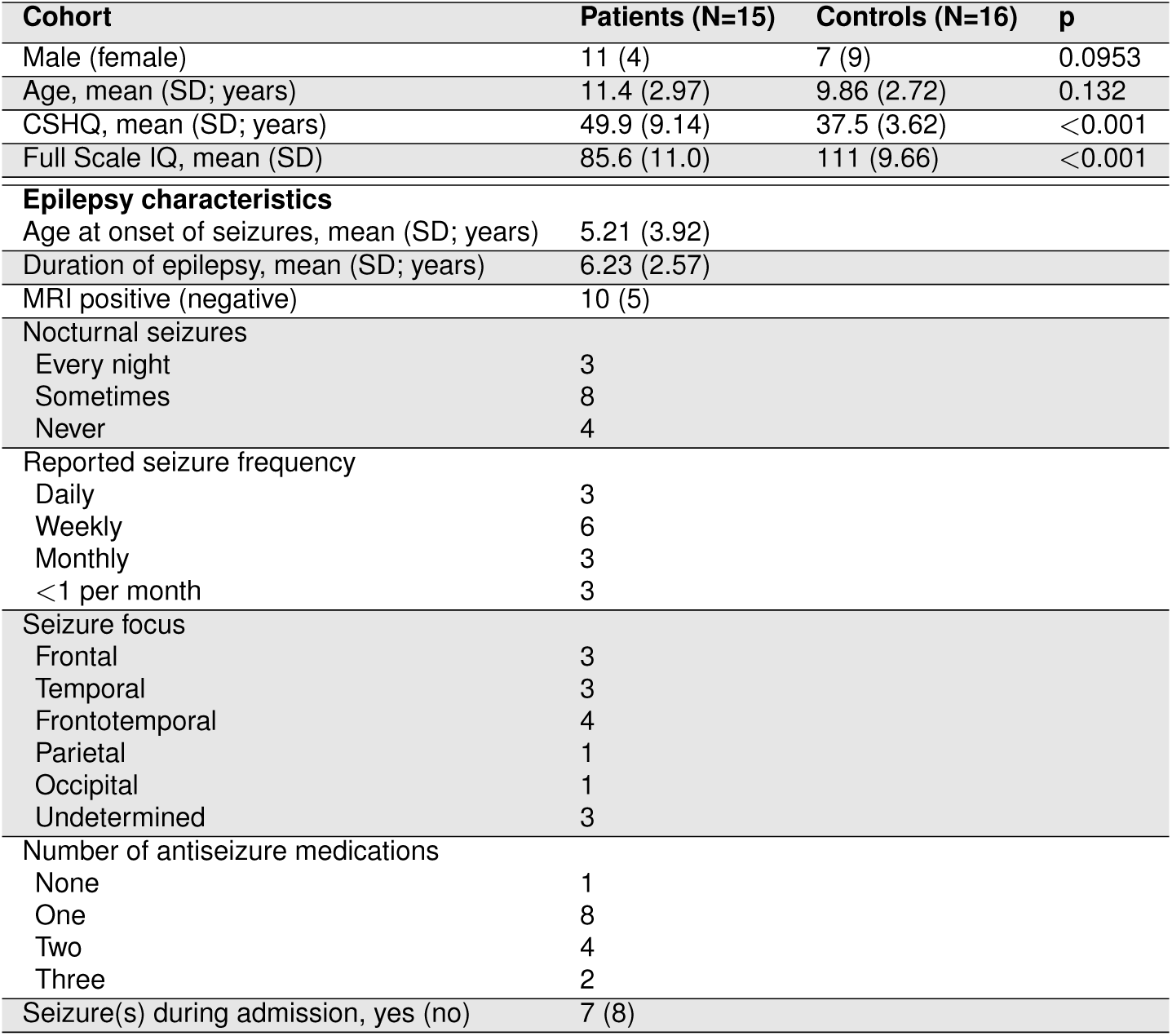
Population demographics. P-values were generated from independent samples t-tests for continuous variables and chi-square tests for categorical variables.

### 5.2. Normalised power spectra in controls and patients

Spectral analysis of the normalised EEG revealed that patients had less relative power in the higher delta range (1.5Hz-4Hz) than controls (Figure 2A). Patients who had a seizure during their hospital admission (referred to herein as ‘patients with seizures’) had even less power in this range than those who did not (referred to herein as ‘patients without seizures’); Figure 2B. The mean and standard error (SE) normalised power from model simulations are shown in Figures 2C-D. It can be seen that the simulations qualitatively recreate the normalised power of the data and the difference observed between each of the groups. Furthermore, for an example control and patient subject, normalised power and time series from the model output are given in Supplemental 4.

**Figure 2:**
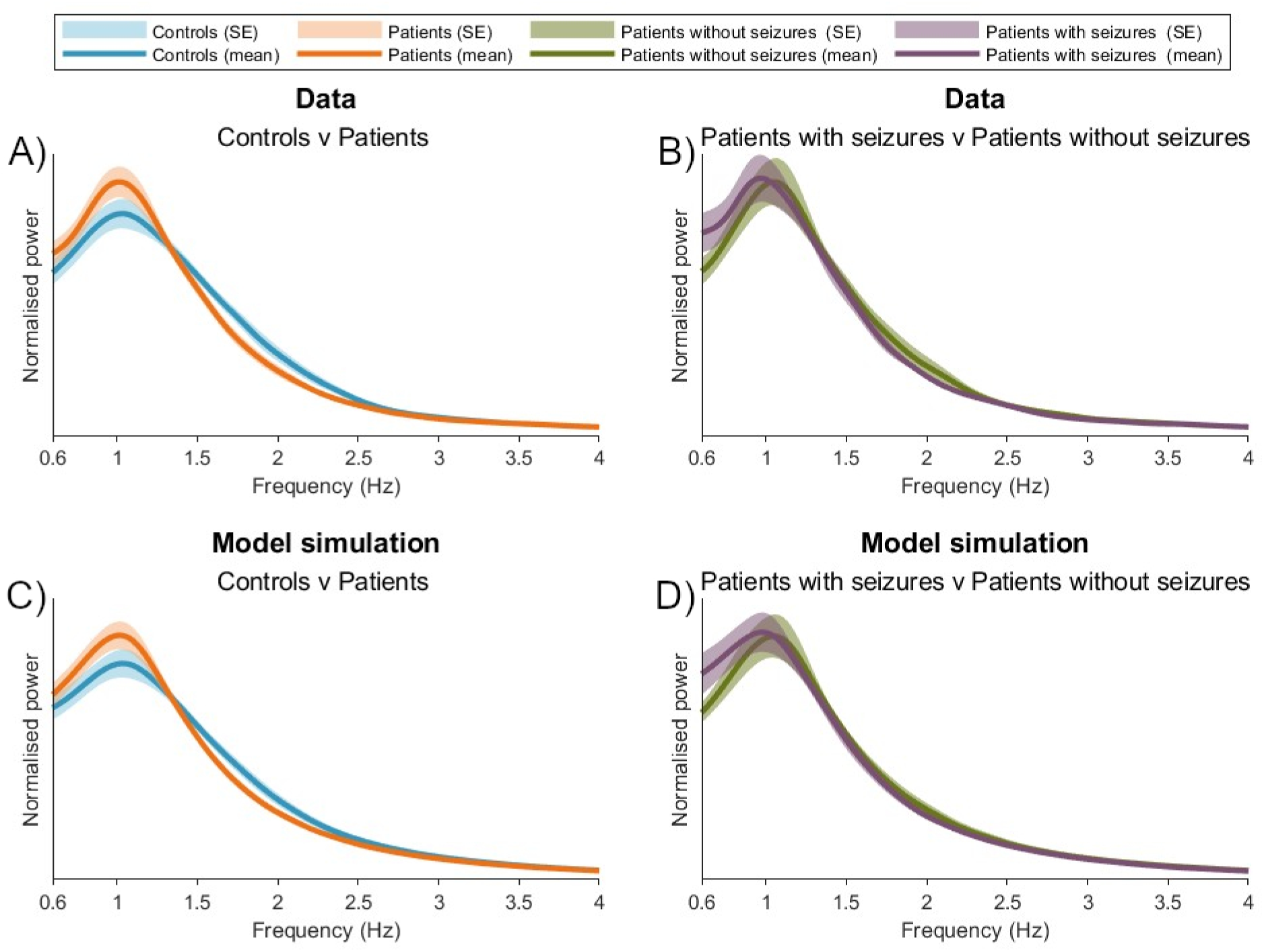
A) Normalised power from control and patient data (mean and SE across subjects). B) Normalised power across patients after splitting patients who had a seizure during their admission (patients with seizures) and patients who did not have a seizure during their admission (patients without seizures). C-D) Shows the same as A-B), but for model simulations after optimising the dynamics to data. Differences in the mean and SE normalised power between groups were recapitulated in the model simulations.

### 5.3. Comparison of model simulations recovered from controls and patients

Parameter distributions recovered from optimising to control and patient data are shown in Supplemental Figure 5. Significant differences were found in synaptic time scales (*γ_e_* and *γ_i_*), excitatory connectivity to excitatory and inhibitory neuronal populations (*N_ee_* and *N_ei_*), firing rate thresholds (*θ_e_* and *θ_i_*) and the excitatory synaptic reversal potential (*E_e_*). These parameters contribute to differences in the emergent properties of the membrane potential and excitability. To interpret the differences observed, we analysed the mean firing rates and mean synaptic currents obtained from the model simulations that best fit the data (Figure 3). It can be seen that patients, and specifically patients who had seizures during their admission, had higher mean firing rates in the excitatory and inhibitory neuronal populations (Figure 3A and B). Patients had significantly larger excitatory synaptic currents onto the excitatory neuronal population than controls (Figure 3C). These differences were driven by the subset of patients who had a seizure during their admittance. Patients with seizures also displayed significantly larger (more negative) inhibitory synaptic currents onto excitatory neurons than controls (Figure 3D). We note that no significant differences were observed in the leak and KNa synaptic currents on the excitatory neuronal populations (Supplemental Figure 6), nor any of the synaptic currents on the inhibitory neuronal populations (Supplemental Figure 7).

**Figure 3:**
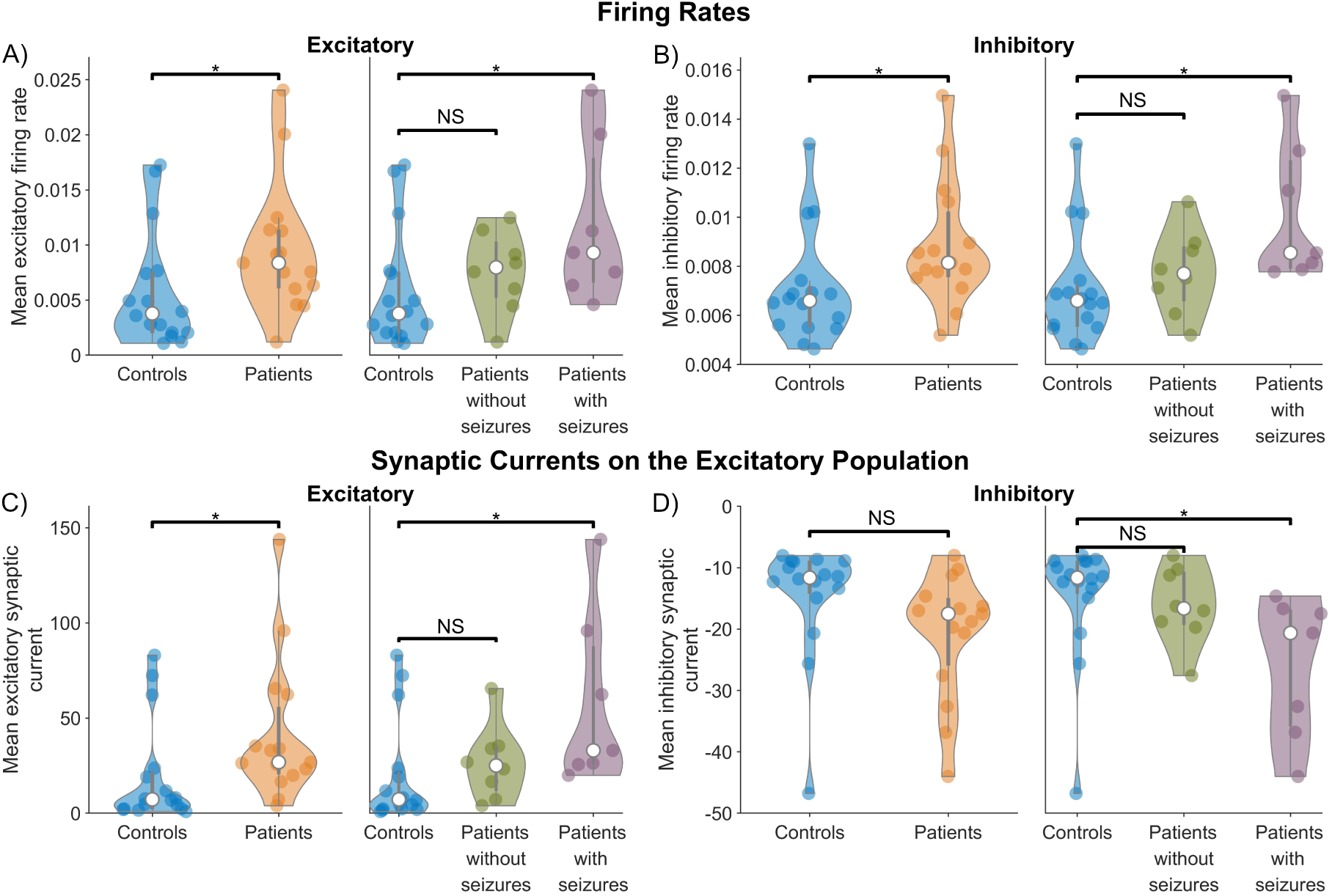
A) Excitatory and B) inhibitory mean firing rates obtained from model simulations. Patients had greater mean excitatory and inhibitory firing rates than controls, as did the subset of patients with seizures during their admission. The mean firing rate of patients without seizures during their admission was not significantly different to controls. C) Excitatory and D) inhibitory synaptic currents on the excitatory neuronal population, obtained from model simulations. In each case, each point on a violin plot gives the mean value of a subject. Using a Mann-Whitney U test, * *p <* 0.05 after Bonferroni correction, NS = not significant.

### 5.4. In silico predictions for intervention

To further understand the influence that each of the synaptic currents has on the power spectrum, we adjusted the conductance of each synapse and simulated the resulting power spectrum. We adjusted each conductance by 40, 50 or 60 percent of its bound (see Supplemental Table 1), starting from the baseline recovered from simulating patient EEG. Note that the parameters governing the excitatory and leak synaptic conductance were reduced, whilst the parameters governing the inhibitory and KNa synaptic conductance were increased, in line with the trends observed in Figure 3. Also note that whereas the results of Figure 3 were specific to synaptic currents on the excitatory neuronal population, here we adjust the excitatory synaptic conductance in general. This allows for the simulation of the effects that drugs (such as antiseizure medications) have on the power spectrum. Figure 4 shows that reducing the excitatory synaptic conductance produced the largest change in the power spectrum. In fact, a 50 percent reduction in the excitatory conductance was sufficient to shift the mean patient power spectrum towards the mean control power spectrum, simulating a “normalisation” of the sleep dynamics of patients (see Figures 4A and 4E). Moreover, the sensitivity of the power spectrum to changes in synaptic conductance was found to be significantly higher for the excitatory conductance than all of the other conductances in the model (Figure 4F-H).

**Figure 4:**
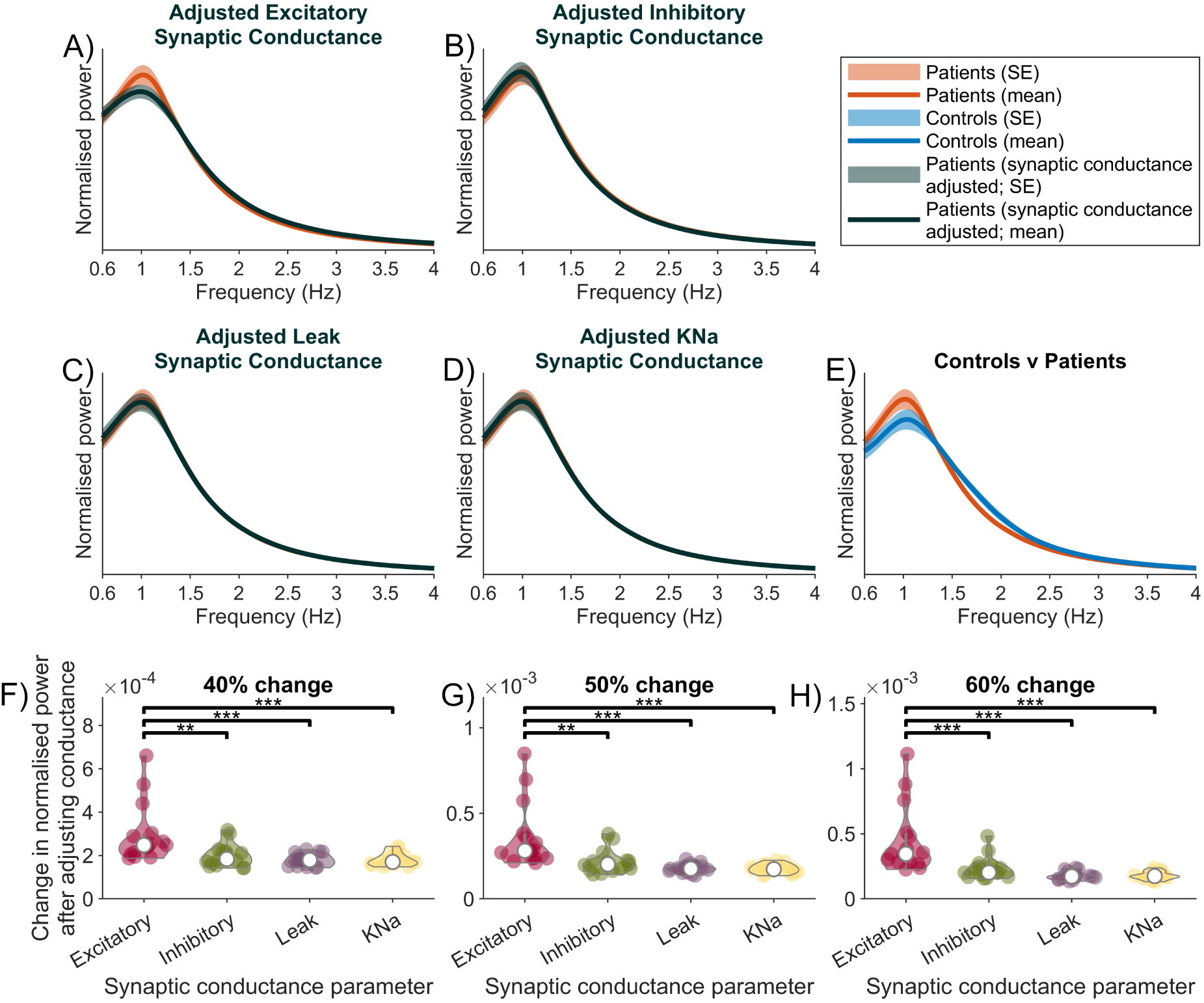
A-D) Normalised power spectra of model simulations from all patients, along with the normalised power spectra obtained after adjusting the excitatory, inhibitory, leak and KNa synaptic conductances by 50% of their range, respectively. E) Normalised power spectra of recorded control and patient EEG, for reference. F-H) Change in normalised power spectra after adjusting the synaptic conductance parameter for 40%, 50% and 60% of the specified parameter bounds (see Supplemental Table 1), respectively. Using a Mann-Whitney U test, \**p <* 0.05, \*\**p <* 0.01 and \*\*\**p <* 0.001.

Hitherto, we used the model to reveal hidden firing rates and synaptic dynamics underlying SWA. We found that alterations to the interactions between excitatory and inhibitory neuronal populations could be revealed from this resting EEG, which was free of overt pathological rhythms. We next sought to understand how these differences might be related to the generation of seizures, which is the defining characteristic of the patient group. To do this, we performed another *in silico* experiment, matching the model output to a prototypic seizure dynamic, the spike-wave discharge (SWD). Figure 5A shows a 2.5s data segment during a SWD, along with a typical model simulation generated using parameters that were recovered by fitting the model to this rhythm (henceforth called SWD models). We compared the mean excitatory synaptic currents of SWD models to patient and control models (which were inferred from resting SWA). We observed that, compared to controls, patients (and in particular the subset of patients that had a seizure during their admission) had excitatory synaptic currents more similar in magnitude to the mean excitatory synaptic currents recovered from SWD models (Figure 5B). This implies that smaller changes to excitatory synaptic currents would cause patient models to generate SWDs, compared to controls. Hence, the models inferred only from SWA dynamics revealed a hidden ictogenicity, as well as a mechanistic explanation (enhanced excitatory synaptic currents and excitability).

**Figure 5:**
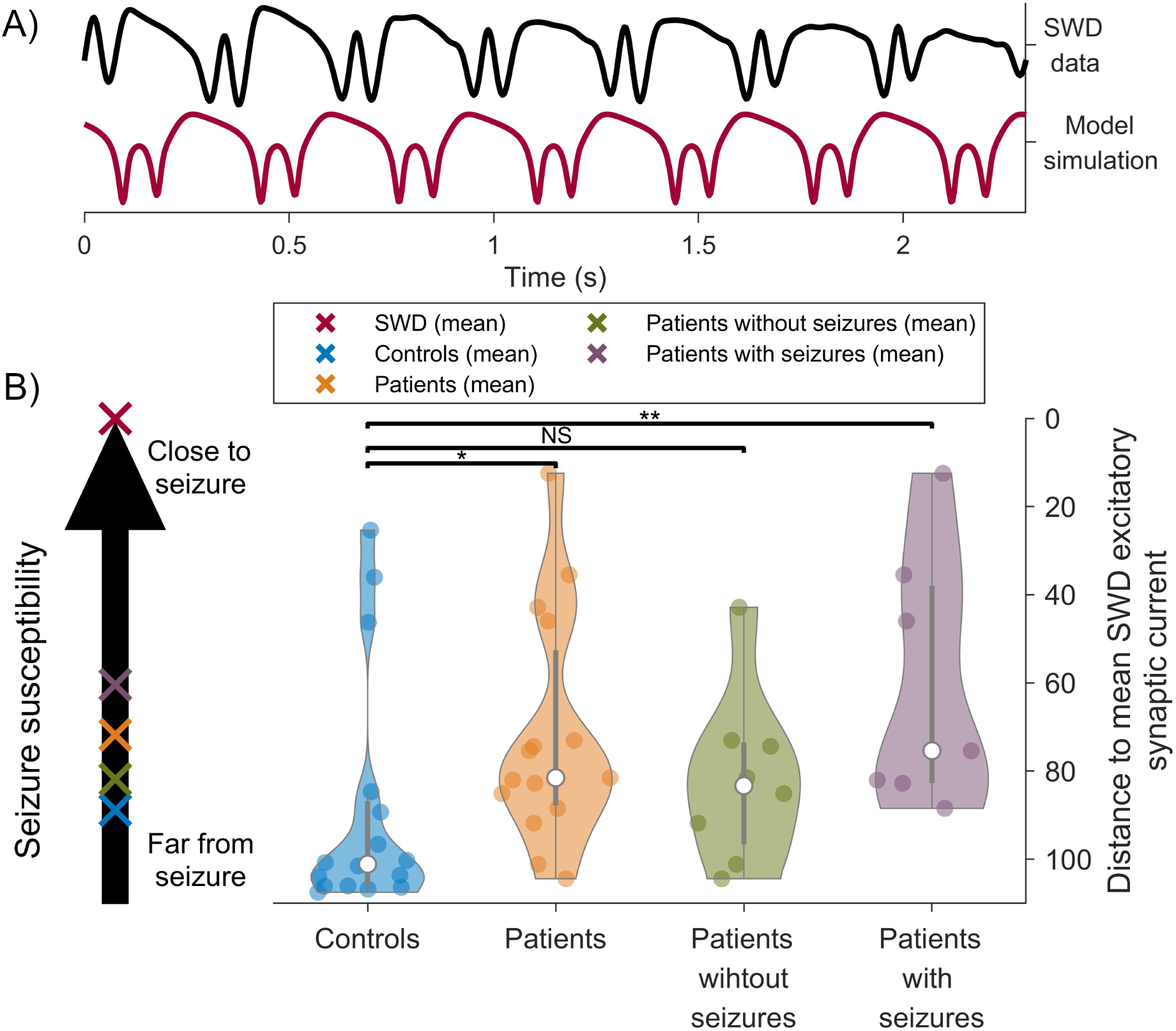
A) SWD data segment and example model simulation obtained after optimising the model dynamics to SWD data. B) Comparison of the absolute difference in excitatory synaptic current obtained from control and patient models (inferred from SWA), compared to the excitatory synaptic current from SWD models. This formed a continuum of seizure susceptibility, as shown on the left of B). Patients, and in particular the subset of patients that had a seizure during their admittance, had excitatory synaptic current more similar to SWD models than controls. Using a Mann-Whitney U test, \**p <* 0.05 and \*\**p <* 0.01 and NS = not significant.

## 6. Discussion

We have used a neural mass model to recapitulate the EEG during deep sleep in children with epilepsy and age-matched healthy controls. These models encapsulate essential features of the system of interest and, by optimising their parameters to data, allow for inferences to be made about the mechanisms that are altered in disease. In the cohort studied, optimising the model parameters for each subject revealed that the differences observed in sleep homeostasis ([7]) at the group level were explained by hyperexcitability in patients, driven predominantly by greater firing rates and excitatory synaptic currents. This indicates that mechanisms can be revealed by neural mass models, and demonstrates that sleep could be a critical paradigm in which mechanistic differences in the epileptic brain manifest during a resting state. Additionally, by comparing the excitatory synaptic current obtained from the resting sleep data with the excitatory synaptic current obtained from simulating a spike-wave discharge, we showed that patients were closer to a seizure state than controls. That is, a smaller adjustment to the patient model could lead them to have seizures *in silico*. These differences were exacerbated in patients who had a seizure within 72 hours after the recording. Thus the propensity of the brain to generate seizures, which is hidden from a visual analysis of the EEG, can be revealed by mathematical models. Crucially, by modifying the parameters governing various channel conductances in the model, we were able to simulate the effects of clinical intervention.

### 6.1. Limitations

Limitations of our study include the small sample size and retrospective sample. Conversely, this meant that all seizures had been recorded as part of continuous study with the period of deep sleep used for analysis. The age range of subjects spanned the developmental period where SWA peaks then falls [36, 37]. We minimised bias by ensuring that the control and patient groups were matched for age, though it is not possible to exclude that delays in the maturation of sleep homeostasis unrelated to seizure burden may have contributed to the observed differences.

### 6.2. Digital twinning to facilitate personalised antiseizure management

EEG remains the key investigation for the diagnosis and monitoring of epilepsy, with interpretation in the clinical setting by visual pattern recognition. However, recent advances in technology are improving biomarkers towards the goals of seizure localisation [38] and forecasting [39]. A further important question concerns the efficacy of treatment; a daily dilemma facing the clinician is how to select the best treatment for a specific patient, which can be conceptualised as an n-of-1 trial.

Building virtual simulations and integrating them with data to make inferences about clinical operations has been termed ‘digital twinning’ [40]. Applying the modelling framework presented, the parameters optimised for each subject could serve as an *in silico* ‘twin’ on which pharmacological, as well as non-pharmacological, interventions could be trialled virtually to aid clinical decision-making. As an example, the findings at the group level from this cohort suggest that utilising antiseizure medication that specifically modulates excitatory synapses, such as perampanel (a known AMPA receptor antagonist [41, 35]), could be beneficial for seizure control in children with focal lesional epilepsies. Further work will include assessing the use of the model on prospective data to see if it can predict the effects of interventions with known mechanisms. In this approach, within-patient comparisons would counter bias due to variations in the maturity of SWA dynamics for age.

### 6.3. Perspectives for guiding stimulation protocols

A further promising area of research concerns the manipulation of slow waves using targeted stimulation protocols. This approach has been implemented through various non-invasive methods, such as transcranial magnetic [42], transcranial direct current [43] and acoustic [44] stimulation. Deep brain stimulation could also be used with enhancement of SWA as a target outcome [45]. Studies exploring stimulation to modulate activity have shown varied levels of success, from no systematic effect on the occurrence of spike-wave activity [46] to some significant improvements in patients with benign epilepsy [47]. In future, stimulation protocols could be tested by implementing them in a mathematical model, such as the one discussed herein. Using a model to help systematize stimulation by guiding the optimal parameters required could be crucial to help improve success rates for the non-pharmacological treatment of epilepsy.

## 7. Conclusions

By recovering parameters of neural mass models from data, we demonstrate the feasibility of using mathematical models to elucidate mechanisms which may contribute to seizure burden in patients with epilepsy. Our results suggest that hyperexcitability underpins the discrepancies observed between children with epilepsy and healthy controls during slow-wave sleep. Furthermore, by simulating seizures in the model, we provide evidence that the observed differences in this resting state may have a causative association with seizure propensity. This approach could generate new biomarkers for seizure susceptibility. Finally, by adjusting synaptic conductances *in silico*, we demonstrate a proof-of-concept for using mathematical models to hypothesise about the most efficacious interventions needed to rectify differences observed on the EEG, with the ultimate goal of improving patient outcomes.

## Data Availability

The data that support the findings of this study are available on request from the corresponding author. The code to generate the figures in the manuscript is publicly available and maintained as a GitHub repository (https://github.com/domdunstan/Neural_Mass_Modelling_Slow_Wave_Sleep_Dunstan_et_al_2024).

## References

[1] Contreras D, Steriade M. Cellular basis of EEG slow rhythms: a study of dynamic corticothalamic relationships. J Neurosci. 1995;15(1):604–622.

[2] Borbély AA. A two process model of sleep regulation. Hum Neurobiol. 1982;1(3):195–204.

[3] Tononi G, Cirelli C. Sleep and the price of plasticity: from synaptic and cellular homeostasis to memory consolidation and integration. Neuron. 2014;81(1):12–34.

[4] Huber R, Mäki H, Rosanova M, Casarotto S, Canali P, Casali A, et al. Human cortical excitability increases with time awake. Cereb Cortex. 2013;23(2):1–7.

[5] de Vivo L, Bellesi M, Marshall W, Bushong E, Ellisman M, Tononi G, et al. Ultrastructural evidence for synaptic scaling across the wake/sleep cycle. Science. 2017;355(6324):507–510.

[6] Halász P, Sźúcs A. Sleep and Epilepsy Link by Plasticity. Front Neurol. 2020;11(911).

[7] Eriksson MH, Baldeweg T, Pressler R, Boyd S, Huber R, Cross J, et al. Sleep homeostasis, seizures, and cognition in children with focal epilepsy. Dev Med Child Neurol. 2023;65(5):701–711.

[8] Vogels TP, Rajan K, Abbott LF. NEURAL NETWORK DYNAMICS. Annu Rev Neurosci. 2005;28:357– 376.

[9] Breakspear M. Dynamic models of large-scale brain activity. Nat Neurosci. 2017;20:340–352.

[10] Deco G, Jirsa VK, Robinson PA, Breakspear M, Friston K. The dynamic brain: from spiking neurons to neural masses and cortical fields. PLoS Comput Biol. 2008;4(8):e1000092.

[11] Freeman WJ. In: Mass Action in the Nervous System. New York: Academic Press; 1975.

[12] Pinotsis DA, Moran RJ, Friston KJ. Dynamic causal modeling with neural fields. Neuroimage. 2012;59(2):1261–1274.

[13] Goodfellow M, Schindler K, Baier G. Intermittent spike–wave dynamics in a heterogeneous, spatially extended neural mass model. NeuroImage. 2011;55(3):920–932.

[14] Wendling F, Bartolomei F, Bellanger JJ, Chauvel P. Epileptic fast activity can be explained by a model of impaired GABAergic dendritic inhibition. Eur J Neurosci. 2002;15(9):1499–1508.

[15] David O, Kiebel SJ, Harrison LM, Mattout J, Kilner JM, Friston KJ. Dynamic causal modeling of evoked responses in EEG and MEG. Neuroimage. 2006;30(4):1255–1272.

[16] West TO, Berthouze L, Farmer SF, Cagnan H, Litvak V. Inference of brain networks with approximate Bayesian computation – assessing face validity with an example application in Parkinsonism. Neuroimage. 2021;236(10):118020.

[17] Lehnertz K. Epilepsy and nonlinear dynamics. J Biol Phys. 2008;34(3):253–266.

[18] Dunstan DM, Richardson MP, Abela E, Akman OE, Goodfellow M. Global nonlinear approach for mapping parameters of neural mass models. PLoS Comput Biol. 2023;19(3):e1010985.

[19] Weigenand A, Schellenberger Costa M, Ngo HV, Claussen JC, Martinetz T. Characterization of K-complexes and slow wave activity in a neural mass model. PLoS Comput Biol. 2014;10(11):e1003923.

[20] Chan S, Pressler R, Boyd SG, Baldeweg T, Cross JH. Does sleep benefit memory consolidation in children with focal epilepsy? Epilepsia. 2017;58(3):456–466.

[21] Owens JA, Spirito A, McGuinn M. The Children’s Sleep Habits Questionnaire (CSHQ): psychometric properties of a survey instrument for school-aged children. Sleep. 2000;23(8):1043–1051.

[22] Lopes da Silva FH, Hoeks A, Smits H, Zetterberg LH. Model of brain rhythmic activity. The alpha-rhythm of the thalamus. Biol Cybern. 1974;15(1):27–37.

[23] Bischoff U, Vogel W, Safronov BV. Na^+^-activated K^+^ channels in small dorsal root ganglion neurones of rat. J Physiol. 1998;510(3):743–754.

[24] MATLAB. Version 9.12.0.1927505 (R2023a). Natick, Massachusetts: The MathWorks Inc.; 2023.

[25] Luque B, Lacasa L, Ballesteros F, Luque J. Horizontal visibility graphs: Exact results for random time series. Phys Rev E. 2009; p. 046103.

[26] Schindler K, Rummel C, Andrzejak RG, Goodfellow M, Zubler F, Abela E, et al. Ictal time-irreversible intracranial EEG signals as markers of the epileptogenic zone. Clin Neurophysiol. 2016;127(9):3051– 3058.

[27] Rugg-Gunn FJ, Stapley HB. In: FROM BENCH TO BEDSIDE. A PRACTICAL GUIDE TO EPILEPSY. ILAE British Branch; 2019.

[28] Bazhenov M, Timofeev I, Fröhlich F, Sejnowski TJ. Cellular and network mechanisms of electrographic seizures. Drug Discov Today Dis Models. 2008;5(1):45–57.

[29] Holmes GL. Benign Focal Epilepsies of Childhood. Epilepsia. 1993;34(S3):S49–S61.

[30] Panayiotopoulos CP. In: A Clinical Guide to Epileptic Syndromes and Their Treatment. Springer Healthcare; 2010.

[31] Van Bogaert P. Epileptic encephalopathy with continuous spike-waves during slow-wave sleep including Landau-Kleffner syndrome. Handb Clin Neurol. 2013;111:635–640.

[32] Steriade M, Contreras D. Spike-Wave Complexes and Fast Components of Cortically Generated Seizures. I. Role of Neocortex and Thalamus. J Neurophysiol. 1998;80(3):1439–1455.

[33] Sills GJ. In: Mechanisms of action of antiepileptic drugs. ILAE (UK Chapter) and the National Society for Epilepsy; 2011.

[34] Eghbali M, Curmi JP, Birnir B, Gage PW. Hippocampal GABA_A_ channel conductance increased by diazepam. Nature. 1997;388(6637):71–75.

[35] Brito da Silva A, Pennifold J, Henley B, Chatterjee K, Bateman D, Whittaker RW, et al. The AMPA receptor antagonist perampanel suppresses epileptic activity in human focal cortical dysplasia. Epilepsia Open. 2022;7(2):488–495.

[36] Ringli M, Huber R. Developmental aspects of sleep slow waves: linking sleep, brain maturation and behavior. Prog Brain Res. 2011;193:63–82.

[37] Chan YS. Sleep architecture and homeostasis in children with epilepsy: a neurodevelopmental perspective. Dev Med Child Neurol. 2020;62(4):426–433.

[38] Ramantani G, Westover MB, Gliske S, Sarnthein J, Sarma S, Wang Y, et al. Passive and active markers of cortical excitability in epilepsy. Epilepsia. 2023;64(S3):S25–S36.

[39] Dell KL, Payne DE, Kremen V, Maturana MI, Gerla V, Nejedly P, et al. Seizure likelihood varies with day-to-day variations in sleep duration in patients with refractory focal epilepsy: A longitudinal electroencephalography investigation. EClinicalMedicine. 2021;37(100):100934.

[40] Erol T, Mendi AF, Doğan D. Digital Transformation Revolution with Digital Twin Technology. 4th International Symposium on Multidisciplinary Studies and Innovative Technologies (ISMSIT). 2020; p. 1–7.

[41] Steinhoff BJ. Introduction: Perampanel—New mode of action and new option for patients with epilepsy. Epilepsia. 2014;55(s1):1–2.

[42] Massimin M, Ferrarelli F, Esser SK, Riedner BA, Huber R, Murphy M, et al. Triggering sleep slow waves by transcranial magnetic stimulation. Proc Natl Acad Sci. 2007;104(20):8496–8501.

[43] Marshall L, Helgadóttir H, Mölle M, Born J. Boosting slow oscillations during sleep potentiates memory. Nature. 2006;444(7119):610–613.

[44] Ngo HVV, Martinetz T, Born J, Mölle M. Auditory closed-loop stimulation of the sleep slow oscillation enhances memory. Neuron. 2013;78(3):545–553.

[45] Buenzli JC, Werth E, Baumann CR, Belvedere A, Renzel R, Stieglitz LH, et al. Deep brain stimulation of the anterior nucleus of the thalamus increases slow wave activity in non-rapid eye movement sleep. Epilepsia. 2023;64(8):2044–2055.

[46] Fattinger S, Heinzle BB, Ramantani G, Abela L, Schmitt B, Huber R. Closed-Loop Acoustic Stimulation During Sleep in Children With Epilepsy: A Hypothesis-Driven Novel Approach to Interact With Spike-Wave Activity and Pilot Data Assessing Feasibility. Front Hum Neurosci. 2019;13:166.

[47] Klinzing JG, Tashiro L, Ruf S, Wolff M, Born J, Ngo HVV. Auditory stimulation during sleep suppresses spike activity in benign epilepsy with centrotemporal spikes. Cell Rep Med. 2021;2(11):100432.

