## Supplemental Material for "Neural mass modelling reveals that hyperexcitability underpins slow-wave sleep changes in children with epilepsy"

*Supplemental 1: Neural Mass Model*

Neural mass models (NMMs) represent the mean macroscopic behaviour of populations of neurons. In this work, we employ the NMM developed by [1]. This model consists of interacting excitatory and inhibitory neuronal populations and aims to recreate activity observed on the EEG during non-REM sleep.

The model dynamics are governed by differential equations that describe the change in activity (membrane potentials and synaptic dynamics) over time. Two main operators are used to generate the dynamics. The first transforms the average density of presynaptic input into a postsynaptic membrane potential. This is modelled by convoluting an impulse response function with the arriving input. The impulse response function used is given by the term $\alpha_j(t) = \gamma_j^2 t \exp(-\gamma_j t)$, for $j = e, i$ representing excitatory and inhibitory synapses, respectively. The second operator transforms the average membrane potential of a population into an outgoing firing rate. This is modelled by a nonlinear sigmoidal function, $S_j(V_j) = \frac{Q_j^{max}}{(1+exp(-\frac{\pi}{3}(V_k-\theta_j)/\sigma_j)}$, for $j = e, i$ representing the excitatory and inhibitory firing rate functions, respectively. An example of an impulse response function and sigmoidal firing rate function is given in Supplemental Figure 1. In full, the model can be expressed as the following set of first and second-order differential equations:

$$\tau_e \frac{dV_e(t)}{dt} = -I_L^e(t) - s_{ee}(t)I_{ee}(t) - s_{ie}(t)I_{ie}(t) - \tau_e C_m^{-1} I_{KNa}(t), \quad (1)$$

$$\tau_i \frac{dV_i(t)}{dt} = -I_L^i(t) - s_{ei}(t)I_{ei}(t) - s_{ii(t)}I_{ii}(t), \quad (2)$$

$$\frac{d^2 s_{ee}(t)}{dt^2} = \gamma_e^2(N_{ee}S_e(V_e(t)) - s_{ee}(t) + \phi(t)) - 2\gamma_e \frac{ds_{ee}(t)}{dt}, \quad (3)$$

$$\frac{d^2 s_{ei}(t)}{dt^2} = \gamma_e^2(N_{ei}S_e(V_e(t)) - s_{ei}(t)) - 2\gamma_e \frac{ds_{ei}(t)}{dt}, \quad (4)$$

$$\frac{d^2 s_{ie}(t)}{dt^2} = \gamma_i^2(N_{ie}S_i(V_i(t)) - s_{ie}(t)) - 2\gamma_i \frac{ds_{ie}(t)}{dt}, \quad (5)$$

$$\frac{d^2 s_{ii}(t)}{dt^2} = \gamma_i^2(N_{ii}S_i(V_i(t)) - s_{ii}(t)) - 2\gamma_i \frac{ds_{ii}(t)}{dt}, \quad (6)$$

$$\tau_{Na} \frac{dNa(t)}{dt} = \alpha_{Na} S_e(V_e(t)) - R_{pump}\left(\frac{Na(t)^3}{Na(t)^3 + K_p^3} - \frac{Na_{eq}^3}{Na_{eq}^3 + K_p^3}\right). \quad (7)$$

The term $-I_L^j = -g_L(V_j(t) - E_L^j)$ represents the passive leak current, with active currents given by, $-s_{jh}(t)I_{jh}(t) = -s_{jh}(t)g_j(V_h(t) - E_j)$ and $-I_{KNa}(t) = -g_{KNa}\frac{0.37}{1+(EC/Na(t))^{nH}}(V_e(t) - E_K)$, with $j = e, i$ and $h = e, i$. As described in [1, 2], these currents incorporate that the postsynaptic current flow will be dependent on the membrane voltage and synaptic reversal potential. In particular, the KNa current was introduced for modelling the sleeping cortex [1]. Furthermore, input from external unmodelled brain regions is given by the term $\phi(t) \sim \mathcal{N}(0, \xi)$, and is assumed to project only onto the excitatory population.

In order to numerically solve the model, Equations 1–7 were converted into a set of eleven first-order stochastic differential equations (SDEs). The Euler–Maruyama method was then used to numerically

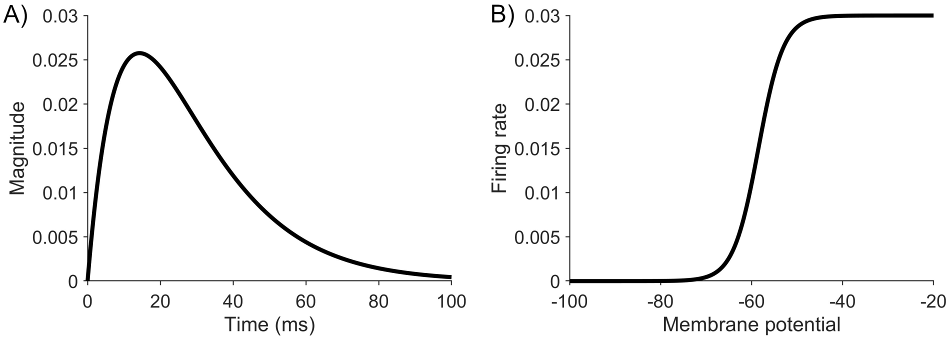


Supplemental Figure 1: A) Example impulse response function used in the NMM. B) Example sigmoidal firing rate function used in the NMM.

solve this set of SDEs, with zero initial conditions. The model was simulated to match the length of the data (30s for slow-wave activity, 2.5s for spike-wave discharge) at a timestep of 0.2ms to ensure accurate convergence. To account for transient dynamics, a further 5s was calculated at the start of the model simulation and then removed. The output of the model was taken to be the membrane potential of the excitatory population, $V_e(t)$. When comparing experimental data and model output, both time series were z-scored. We note that the spike-wave discharge was simulated in the noise-free deterministic case (i.e. $\xi = 0$).

Supplemental Table 1 provides a list of all the parameters in the model, along with a physiological interpretation, typically used values and parameter bounds. In general, these bounds were set to a 25% deviation from the typical values used (see [1] and references therein). This is with the exception of the timescale parameters, in which their bounds were set to 100% of the typical values, and the synaptic reversal potentials for the excitatory and inhibitory populations, which were constructed to ensure that excitatory currents were depolarizing and inhibitory currents were hyperpolarizing (see Supplemental Table 1). Moreover, Supplemental Figure 2 shows the mean excitatory and inhibitory membrane potentials and synaptic currents on the excitatory population obtained from simulations of random parameter values within the bounds specified.

| Parameter | Interpretation | Typical Value | Bounds |
|---|---|---|---|
| $N_{ee}$, $N_{ei}$, $N_{ie}$, $N_{ii}$ | Number of excitatory - excitatory/ excitatory - inhibitory/ inhibitory - excitatory/ inhibitory - inhibitory synaptic connections | 120, 90, 72, 90 | $[90, 150]$, $[67.5, 112.5]$, $[54, 90]$, $[67.5, 112.5]$ |
| $\gamma_e$, $\gamma_i$ | Excitatory/ inhibitory postsynaptic potential rate constant | $0.07/ms$, $0.0586/ms$ | $[0, 0.14]$, $[0, 0.1172]$ |
| $\tau_e, \tau_i$ | Passive excitatory/ inhibitory membrane time constant | $30ms$, $30ms$ | $[22.5, 37.5]$, $[22.5, 37.5]$ |
| $Q_e^{max}$, $Q_i^{max}$ | Maximum mean firing rate of excitatory/ inhibitory population | $0.03/ms$ , $0.06/ms$ | $[0.0225, 0.0375]$, $[0.045, 0.075]$ |
| $\theta_e, \theta_i$ | Excitatory/ inhibitory firing rate threshold | $-58.5mV$, $-58.5mV$ | $[-73.1250, -43.8750]$, $[-73.1250, -43.8750]$ |
| $\sigma_e, \sigma_i$ | Excitatory/ inhibitory firing rate standard deviation | $5mV$, $6mV$ | $[3.75, 6.25]$, $[4.5, 7.5]$ |
| $E_e$, $E_i$ | Excitatory/ inhibitory mean synaptic reversal potential | $0mV$, $-70mV$ | $[-50, 50]$, $[-200, -100]$ |
| $E_L^e$, $E_L^i$ | Excitatory/ inhibitory mean synaptic reversal potential associated with leak channels | $-64mV$, $-64mV$ | $[-80, -48]$, $[-80, -48]$ |
| $g_e$, $g_i$, $g_L$ | Excitatory/inhibitory/leak synaptic conductance | $1mS/cm^2$, $1mS/cm^2$, $1mS/cm^2$ | $[0.75, 1.25]$, $[0.75, 1.25]$, $[0.75, 1.25]$ |
| $C_m$ | Membrane Capacity | $1\mu F/cm^2$ | $[0.75, 1.25]$ |
| $R_{pump}$ | Sodium pump capacity | $0.09mMms^{-1}$ | $[0.0675, 0.1125]$, |
| $\tau_{Na}$ | Sodium time constant | $1.3ms$ | $[0.975, 1.625]$ |
| $\alpha_{Na}$ | Sodium influx | $2mMms$ | $[1.5, 2.5]$ |
| $Na_{eq}$ | Sodium resting state | $9.5mM$ | $[7.125, 11.875]$ |
| $g_k^{Na}$ | Synaptic conductance associated with sodium-potassium pump | $3$ | $[2.25, 3.75]$ |
| $E_k$ | Mean synaptic reversal potential associated with sodium-potassium pump | $-100mV$ | $[-125, -75]$ |
| $EC$ | Sodium for half activation | $38.7$ | $[29.025, 48.375]$ |
| $n_H$ | Hill coefficient | $3.5$ | $[2.625, 4.375]$ |
| $K_p$ | Pump constant | $15$ | $[11.25, 18.75]$ |
| $\xi$ | Standard deviation of noise perturbation | - | $[3.75, 6.25]$ |

Supplemental Table 1: Model parameter values with interpretation, a typically chosen value [1] and parameter bounds.

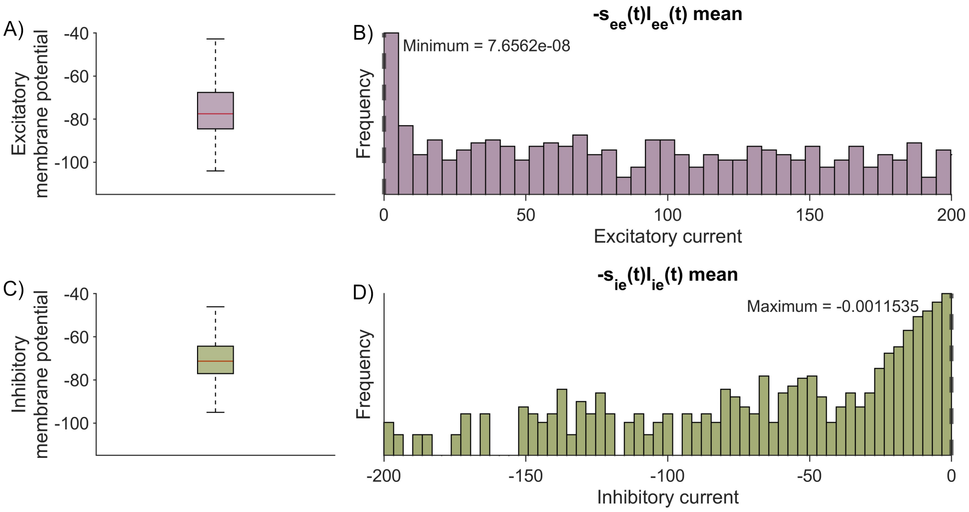


Supplemental Figure 2: A) Mean excitatory membrane potential and B) mean excitatory synaptic currents obtained from simulations of 1000 random parameter sets within the bounds specified in Supplemental Table 1. C) Mean inhibitory membrane potential and D) mean inhibitory synaptic currents obtained from simulations of 1000 random parameter sets within the bounds specified in Supplemental Table 1. Excitatory synaptic currents are exclusively depolarizing within these parameter ranges (minimum current $> 0$) and inhibitory synaptic currents are exclusively hyperpolarizing (maximum current $< 0$).

*Supplemental 2: Multiobjective Optimisation*

In this study, we used a multiobjective evolutionary algorithm to recover parameters from EEG data. In particular, we used the Non-dominated Sorting Genetic Algorithm-II (NSGA-II) [3]. A central reason for using NSGA-II is because of the complex bifurcation structure that is known to exist in an NMM. In general, multiobjecitve optimisation aims to minimise **F(x)** $= (f_1(\mathbf{x}), f_2(\mathbf{x}), ..., f_d(\mathbf{x}))$, given a set of constraints on the inputs **x** $= (x_1, x_2, ..., x_g)$. **F(x)** forms the $d$-dimensional objective space and **x** forms the $g$-dimensional decision space. In this study, the decision space (or model parameter inputs) is bounded by the constraints on parameters (as defined in Supplemental Table 1). Following from [4], we define two objectives (i.e. $d = 2$) to describe the difference between model output and data. The first objective is the sum of squared error between the power spectral density (PSD) from the model simulation and the PSD from the data (after the data was filtered by a 4$^{\text{th}}$ order Butterworth filter between 0.3Hz and 10Hz). This is defined by,

$$Obj_1 = \sum_{\omega_1} (\mu_{data}^{\omega_1} - \mu_{model}^{\omega_1})^2, \tag{8}$$

where $\mu_{data}^{\omega_1}$ gives the data PSD at frequency $\omega_1$ and $\mu_{model}^{\omega_1}$ gives the model PSD at frequency $\omega_1$. In the interest of modelling slow-wave sleep, $\omega_1$ was calculated across 0.6Hz to 10Hz, with a resolution of 0.025Hz.

The second objective used is based on the horizontal visibility graph (HVG) algorithm, which provides a mapping from a time series to an undirected network [5]. Here, each time point becomes a node in the network and an edge is drawn between two nodes in the network if it is possible to trace a horizontal line between the two data points, without intersecting an intermediate point. Formally, for a time series $\{a_i\}_{i=1,...,N}$ of $N$ data points, the HVG algorithm sets nodes $V = \{1, ..., N\}$ and edges between nodes $i$ and $j$ iff $a_i, a_j > a_k \ \forall \ i < k < j$. In this study, we used the extended version of this algorithm known as the weighted HVG, whereby if there exists an edge between two nodes, then that edge is weighted by the difference in amplitude between the time points (latter time point minus former time point). Retaining properties of the weighted HVG in NMM simulations have previously been shown to be useful in terms of refining the plausible dynamics (see [4]). We use the two-sample Kolmogorov-Smirnov test statistic to generate the objective for the difference between the weighted HVG from the model simulation and the data. In particular, for $F : \mathbb{R} \to [0, 1]$ and $G : \mathbb{R} \to [0, 1]$ representing the weighted HVG sum of node weights cumulative distribution function of the data and model simulation respectively, and $y \in \mathbb{R}$, the second objective is defined as,

$$Obj_2 = \sup_y \mid F(y) - G(y) \mid . \tag{9}$$

Due to the stochasticity of the model simulation, and to ensure accurate objective calculations, we simulate the model and calculate each objective function twenty times and then assign the mean value obtained from these repeats as the objective for that parameter set. We note that all parameters were varied in the optimisation, and a fixed population size of 500 and a generation number of 50 were used for accurate convergence.

*Supplemental 3: Epilepsy Characteristics*

Supplemental Table 2 gives details on the individual patient characteristics.

| Patient ID | Sex | Aetiology | Seizure focus | Seizure frequency | Seizure(s) during admission | Antiseizure medication(s) |
|---|---|---|---|---|---|---|
| 1 | F | | Occipital | Weekly | No | Levetiracetam |
| 2 | M | Focal Cortical Dysplasia | Temporal | Monthly | No | Lamotrigine |
| 3 | M | Glioneuronal tumour | Temporal | Monthly | No | Lamotrigine, Sodium valproate |
| 4 | F | MRI negative | Undetermined | Weekly | Yes | Clobazam |
| 5 | M | MRI negative | Frontal | Daily | Yes | Carbamazepine |
| 6 | M | Ganglioglioma | Temporal | Weekly | Yes | Levetiracetam |
| 7 | F | Focal Cortical Dysplasia | Fronto-temporal | Monthly | No | Carbamazepine |
| 8 | F | | Parietal | Weekly | No | None |
| 9 | M | | Fronto-temporal | Daily | Yes | Lamotrigine |
| 10 | M | Focal Cortical Dysplasia | Fronto-temporal | Weekly | Yes | Levetiracetam, Sodium valproate, Oxcarbazepine |
| 11 | M | MRI negative | Fronto-temporal | Weekly | Yes | Carbamazepine, Topiramate |
| 12 | M | | Undetermined | $<$1 per month | No | Levetiracetam, Topiramate, Clobazam |
| 13 | M | MRI negative | Undetermined | $<$1 per month | No | Levetiracetam, Sodium valproate |
| 14 | M | MRI negative | Frontal | Daily | Yes | Lacosamide |
| 15 | M | Focal Cortical Dysplasia | Frontal | $<$1 per month | No | Levetiracetam, Sodium valproate |

Supplemental Table 2: Paitent demographics. Note that age and age at onset have been removed for identification purposes.

*Supplemental 4: Results from comparing control and patient models*

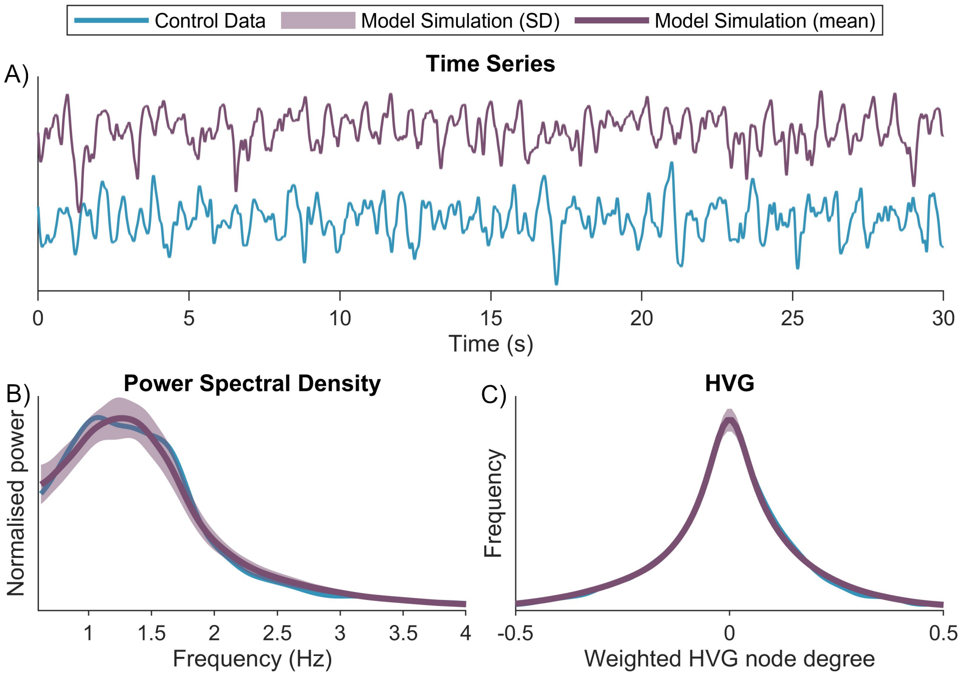


Supplemental Figure 3: A) Example time series, B) normalised power and C) weighted HVG distribution for a control data subject and model output, after optimising the model to data. The example shows the model dynamics closely replicate properties of the EEG data.

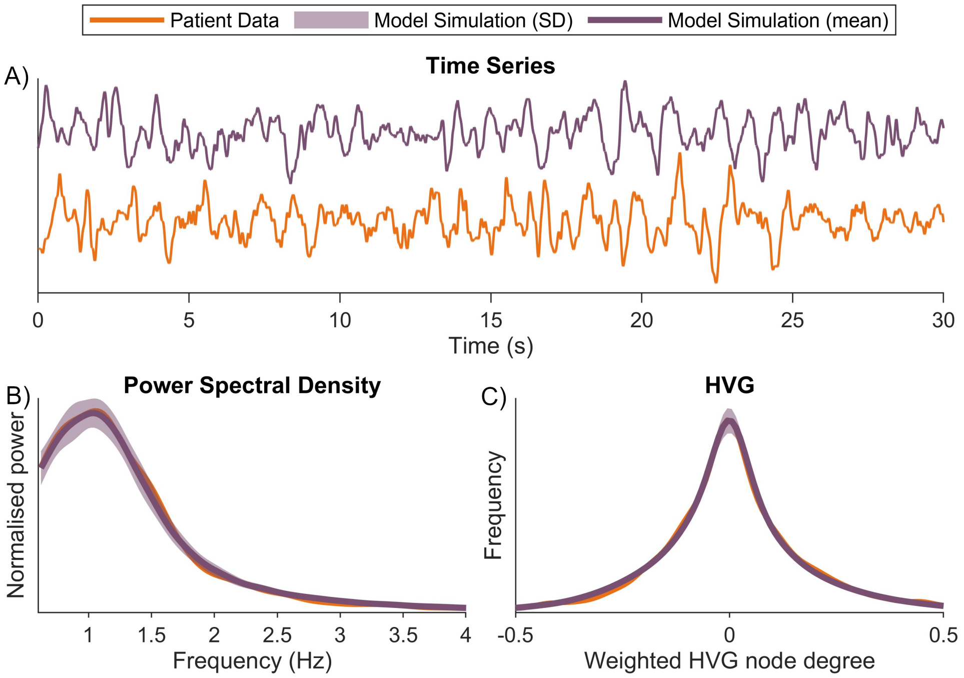


Supplemental Figure 4: A) Example time series, B) normalised power and C) weighted HVG distribution for a patient data subject and model output, after optimising the model to data. The example shows the model dynamics closely replicate properties of the EEG data.

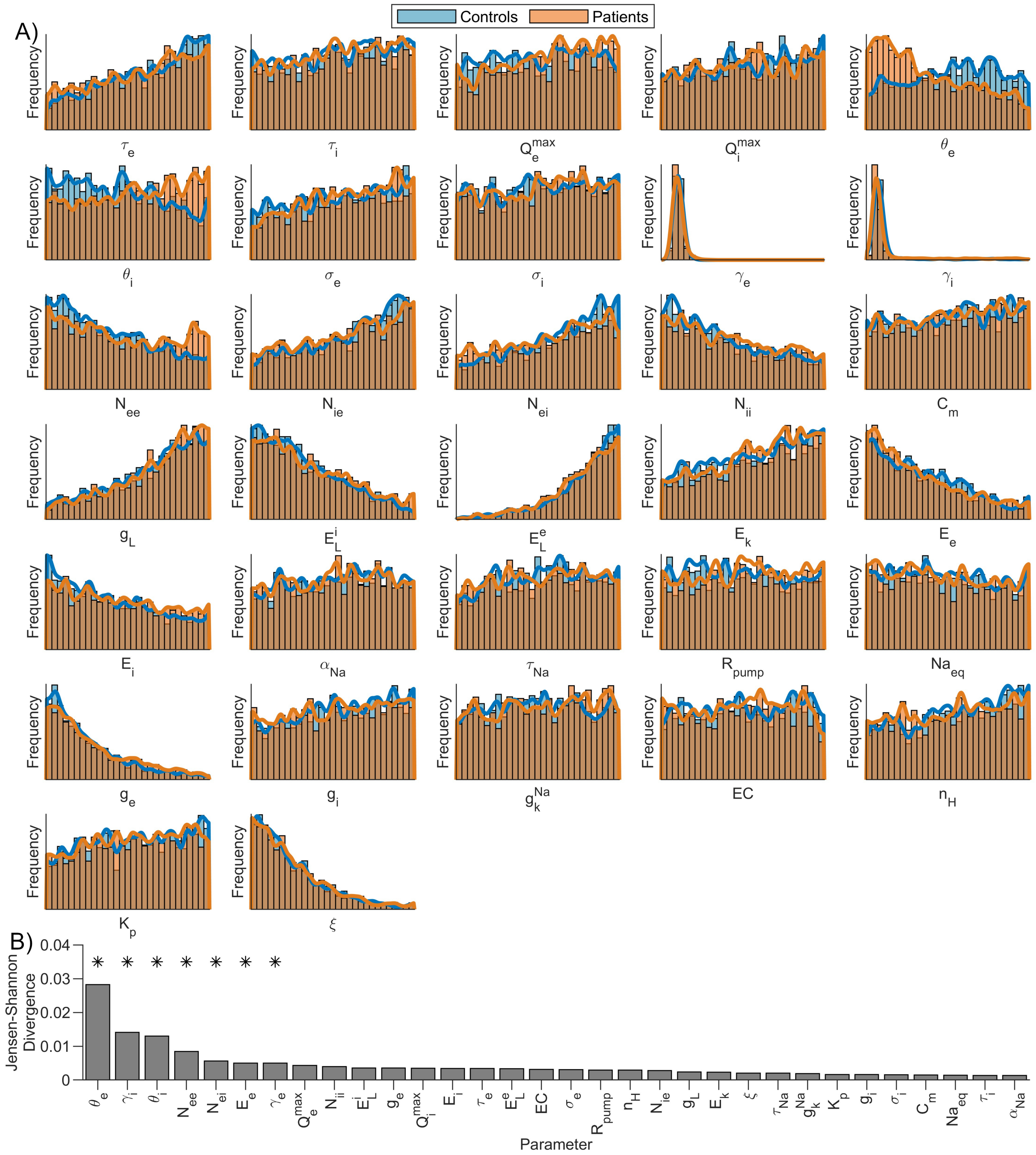


Supplemental Figure 5: Parameter distributions recovered from fitting to all control and patient subjects. A) Univariate parameter distributions. B) Jensen-Shannon Divergence between controls and patients for each parameter. * Gives parameters with significant differences between controls and patients after Bonferroni correction, obtained via bootstrap sampling.

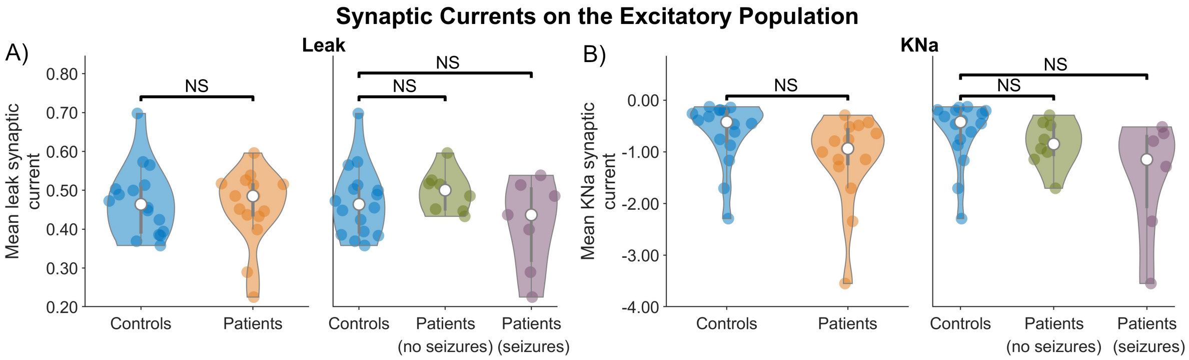


Supplemental Figure 6: A) Leak and B) KNa synaptic currents on the excitatory neuronal population, obtained from model simulations. In each case, each point on a violin plot gives the mean value of a subject. No significant differences were found (NS = not significant) after Bonferroni correction, using a Mann-Whitney U test.

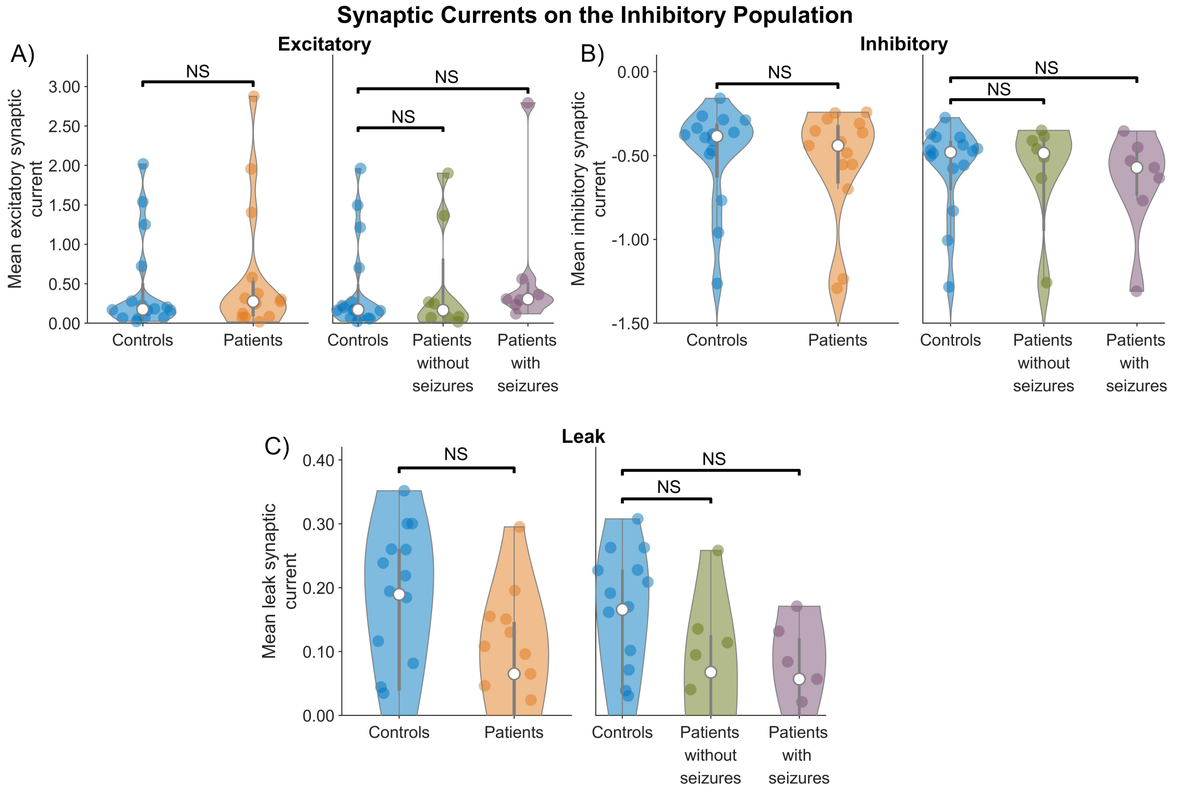


Supplemental Figure 7: A) Excitatory, B) inhibitory and C) leak synaptic currents on the inhibitory neuronal population, obtained from model simulations. In each case, each point on a violin plot gives the mean value of a subject. No significant differences were found (NS = not significant) after Bonferroni correction, using a Mann-Whitney U test.

### References


[1] Weigenand A, Schellenberger Costa M, Ngo HV, Claussen JC, Martinetz T. Characterization of K-complexes and slow wave activity in a neural mass model. PLoS Comput Biol. 2014;**10**(11):e1003923.

[2] Liley DT, J CP, Dafilis MP. A spatially continuous mean field theory of electrocortical activity. Netw Comput Neural Syst. 2002;**13**(1):67–113.

[3] Deb K, Pratap A, Agarwal S, Meyarivan T. A fast and elitist multiobjective genetic algorithm: NSGA-II. IEEE Transactions on Evolutionary Computation. 2002;**6**(2):182–197.

[4] Dunstan DM, Richardson MP, Abela E, Akman OE, Goodfellow M. Global nonlinear approach for mapping parameters of neural mass models. PLoS Comput Biol. 2023;**19**(3):e1010985.

[5] Luque B, Lacasa L, Ballesteros F, Luque J. Horizontal visibility graphs: Exact results for random time series. Phys Rev E. 2009;**80**:046103.
